# The Impact of Universal Transport Media and Viral Transport Media Liquid Samples on a SARS-CoV-2 Rapid Antigen Test

**DOI:** 10.1101/2021.05.12.21257107

**Authors:** J Mayfield, P Hesse, D Ledden

**Affiliations:** Siemens Healthcare Diagnostics Inc., Mishawaka, IN, U.S.; Siemens Healthcare Diagnostics Inc., Marburg, Germany

## Abstract

The impact of universal transport media (UTM) and viral transport media (VTM) liquid samples on the performance of the Healgen Scientific Rapid COVID-19 Antigen Test was investigated. Twelve different UTM/VTM liquid samples were added at different dilutions to the extraction buffer, and 2 of 12 generated false-positive results. To understand the cause of these false-positive results, the effect of extraction buffer dilution on sample pH, surfactant concentration, and ionic strength were investigated. The most important factor in UTM/VTM liquid sample dilution of the extraction buffer was ionic strength as measured by conductivity. Dilutions with conductivity below ∼17 mS/cm can induce a false-positive result. It was also noted that the ionic strength of UTM/VTMs can vary, and those with low ionic strength can be problematic. To rule out the effect of other common components found in UTMs/VTMs, several materials were mixed with extraction buffer and tested at high concentrations. None was shown to produce false-positive results.

## Introduction

The introduction of COVID-19 rapid antigen tests has had a big impact on controlling the spread of SARS-CoV-2 infections during the 2019 pandemic.^1^ Although these tests may be analytically less sensitive than the gold standard RT-PCR assay, they are fast (i.e., 10–20 minutes versus hours to days), much less expensive, and do not require highly trained professionals to perform the testing. Rapid antigen testing requires the acquisition of sample via swab followed by extraction of the viral particles to release the nucleocapsid protein (N-protein) antigen. The extracted sample is then placed in the sample well of the test device, and the results can be detected visually or instrument-read.

Current upper respiratory sample types include nasopharyngeal swabs, nasal swabs, and oropharyngeal swabs. Alternative sample types such as saliva, oral fluid, gargle and wash, and breath are being investigated.^2^ It is preferable that the sample swabs be tested immediately (or within an hour). However, in some cases, particularly for retrospective evaluations of these rapid antigen tests, these swabs, which are also used for RT-PCR testing, are stored in either universal transport medium (UTM) or viral transport medium (VTM). In these cases, the sample swab is placed in 1 to 3 mL of UTM/VTM. These sample types are problematic in that some manufacturers (e.g., Becton Dickinson, Healgen, and Abbott) do not support the use of UTM/VTMs, other manufacturers (e.g., AccessBio, SD Biosensor, and Quidel) recommend specific UTM/VTMs, and some manufacturers (e.g., Quidel) specify that certain UTM/VTM products should not be used.^2-5^

It is important for rapid antigen manufacturers to understand how UTM/VTM samples may affect assay performance.^6^ The major issues with these samples are swab sample dilution (e.g., decreased analyte concentration), swab sample dilution on extraction buffer components (surfactant, pH, and ionic strength), and UTM/VTM formulations (e.g., fetal calf serum, bovine serum albumin, gelatin, amino acids, metabolites, and dyes). To understand how UTM/VTM samples affect the performance of the Healgen rapid antigen test, we performed three different studies: 1) a dilution study using several different UTM/VTMs, 2) a study to determine the impact of UTM/VTM liquid sample dilution on the surfactant concentration of the viral extraction solution, pH, and ionic strength, and 3) a study to investigate concentrations of UTM/VTM components.

## Materials and Methods

In the first study, 12 different UTM/VTMs from seven different vendors (i.e., VRW, COPAN, bioMerieux, Hain Lifescience, BD, Hardy Diagnostics, and Xebios Diagnostics) were acquired and tested. All these materials were diluted with 300 µL of the Healgen extraction buffer at two different dilutions (i.e., 50 µL sample and 300 µL sample). The first dilution (1:6) was chosen because it is known that the swab will likely pick up a 50 µL nasal or oral sample; the second dilution (1:1) was chosen to represent the maximum volume of UTM/VTM used to compensate for the loss of antigen concentration due to sample dilution.

The Coronavirus Ag Rapid Test Cassette (Healgen Scientific Limited Liability Company, Houston TX, USA) lot number used for this study was 2011153, Exp: 2022-10-31.

The second study focused on the impact of liquid VTM/UTM sample dilution on the pH, surfactant concentration, and ionic strength. Extraction buffer mimics were prepared using Tris buffer (Sigma Trizma PreSet Crystals) in ultrapure water (18.2 MΩ-cm at 25°C) at pH values of 7 and 8. In addition, these extraction buffer mimics were prepared with and without 1.0% surfactants (Sigma, Triton X-100, and Tween-20) or with and without 0.2 M NaCl (Sigma). Conductivity and pH measurements were performed on a Metler Toledo Seven Compact Conductivity meter and a Fisher Scientific ACCUMET AE/50 pH meter, respectively. The conductivity meter was calibrated before use with VWR Symphony one-point calibrator at 1412 µS/cM. The pH meter was calibrated before use using multiple levels of Fisher Chemical buffer solutions (pH 4, 7, and 10). The kit extraction buffer, extraction buffer dilutions (saline and water), and extraction buffer mimics were tested neat and at 1:6, 1:4, 1:2, and 1:1 dilutions. In all cases, 100 µL of sample was applied to the sample well of the device via pipette (RAININ single-channel manual pipette, 20–200 µL), and the results were read visually at 15 minutes. Duplicate tests were measured unless indicated.

The third study examined the impact of the different components in the UTM/VTMs (e.g., fetal calf serum, bovine serum albumin, gelatin, amino acids, metabolites, and dyes). Preliminary studies included adding concentrations of BSA (Sigma 100 mg/mL), fetal bovine serum (Sigma, 50%), gelatin (Sigma, 1%), yeast extract (Sigma, 10%), and lactalbumin (Sigma, 100 mg/mL) to the existing extraction buffer. A viral transport media mimic developed according to Centers for Disease Control and Prevention procedure (3) using 1X Hank’s balanced salt solution (HBSS, Sigma) and 2% heat-inactivated fetal bovine serum (FBS, Sigma) in water was prepared and tested similarly to the commercially available VTMs listed above. In all cases, 100 µL of sample was applied to the sample well of the device via pipette, the results were read visually at 15 minutes, and images were recorded. Duplicate tests were measured unless indicated.

The Coronavirus Ag Rapid Test Cassettes (Healgen, Lot 2009087, Exp. 2022-08-31) and CLINITEST® Rapid COVID-19 Antigen Test (manufactured by Healgen Scientific LLC, distributed by Siemens Healthcare Diagnostics Inc., Lot 2010185, Exp. 2022-09-30 and Lot 2010187, Exp. 2022-09-30) were used in the second and third studies.

## Results

Three different studies were performed to determine the impact of UTM/VTM liquid samples on the Healgen Coronavirus Ag Rapid Test. The results of these studies are summarized below.

The first study involved screening several different UTM/VTM vendors’ materials at two different dilutions of extraction buffer to determine the impact on rapid antigen test performance. Twelve different materials from seven different vendors were evaluated. The three materials from bioMerieux were tested only at the 1:6 dilution. Therefore, it is impossible to know if these materials would have generated false-positive results at the 1:1 dilution. In any event, 2 of the 12 materials (i.e., COPAN 144C and Hardy Diagnostics Viral Transport Medium) generated weak false-positive results, as shown in Table 1.

**Table 1.** Universal transport media and viral transport media screened and 1:6 and 1:1 dilution test results.

| Vendor | UTM/VTM | 1:6 Dilution | 1:1 Dilution |
| --- | --- | --- | --- |
| VRW | 710-0438 | Negative | Negative |
| VRW | 710-0442 | Negative | Negative |
| COPAN | 144C | Negative | Weak positive |
| COPAN | 147C | Negative | Negative |
| bioMerieux | PORTAGERM tubes (Port-T) | Negative | Not tested |
| bioMerieux | PORTAGERM bottle (Port-F) | Negative | Not tested |
| bioMerieux | PORTAGERM Armies Agar + Buffer | Negative | Not tested |
| Hain Lifescience | COPAN 350 C | Negative | Negative |
| Hain Lifescience | 140 C | Negative | Negative |
| Becton Dickinson | Universal Viral Transport Medium | Negative | Negative |
| Hardy Diagnostics | Viral Transport Medium | Negative | Weak positive |
| Xebios Diagnostics | GLY 70-2025 | Negative | Negative |

Based on these results, two more studies were performed to determine if weak false-positive results were due to changes in the extraction buffer properties via dilution (e.g., pH, surfactant concentration, or ionic strength) or one or more of the components in the two UTM/VTMs in question.

The importance of surfactant, pH, and ionic strength in the extraction buffer was tested with respect to false positives. Results are shown below in Table 2. Buffer at pH 7 and 8 was prepared with and without the additives. Solutions with both additives yielded negative results at pH 7 and 8. The addition of surfactant without salt resulted in false positives at both pH 7 and 8. The absence of surfactant with salt produced negative test results at both pH 7 and 8. Absence of both additives also produced a positive test line at both pH levels. Solutions without salt added generated false-positive results, suggesting a critical role for ionic strength.

**Table 2.** Extraction buffer components (surfactants, pH, ionic strength) tested for false-positive results.

| Sample | pH | Surfactant | Salt | Result |
| --- | --- | --- | --- | --- |
| Tris buffer | 8.0 | Yes | Yes | Negative |
| Tris buffer | 8.0 | Yes | No | Positive |
| Tris buffer | 8.0 | No | Yes | Negative |
| Tris buffer | 8.0 | No | No | Positive |
| Tris buffer | 7.0 | Yes | Yes | Negative |
| Tris buffer | 7.0 | Yes | No | Positive |
| Tris buffer | 7.0 | No | Yes | Negative |
| Tris buffer | 7.0 | No | No | Positive |

False-positive results due to the absence of sodium chloride suggest that extraction buffer ionic strength measured as conductivity (mS/cm) may play a role in reducing nonspecific interactions. To test this hypothesis, several samples were prepared with conductivity ranging from high to low by mixing assay extraction buffer with ultrapure water (measured conductivity = 0.15 mS/cm) and 0.9% saline (measured conductivity = 17 mS/cm). Volume ratios tested were determined based on potential use cases for samples stored in VTM or buffer. Ultrapure water represents the most extreme case, where the initial sample conductivity is very low. Saline samples represent cases where samples are stored in a higher-conductivity solution. Results of three replicates and two rapid antigen test reagent lots are shown in Tables 3 and 4 below. Samples with conductivity below ∼17.5 mS/cm generated false-positive test results. As expected, mixing extraction buffer (EB) with water, the low-conductivity solution, generated false positives at a dilution of 2:3 (sample:EB). There were small differences between this cutoff for the two lots tested, suggesting some variability. Mixing saline, a higher-conductivity sample, with extraction buffer resulted in no false positives when mixed up to 2:1 (sample:EB). In each ratio, the conductivity of the final solution was greater than 17.5 mS/cm. A false positive occurred only when saline was run without EB, with conductivity at ∼17 mS/cm.

**Table 3.** False positives with water as sample. Results from mixing extraction buffer with varying amounts of water and testing for false positives and conductivity. False positives occurred when the water was mixed with extraction buffer, making the conductivity <17.5 mS/cm.

| Extraction Buffer<br>(μL) | Water<br>(μL) | Lot 2010185/20201307 | | Lot 2010187/20201308 | |
| --- | --- | --- | --- | --- | --- |
|  |  | Conductivity<br>(mS/cm) | False Positive/<br>Replicates | Conductivity<br>(mS/cm) | False Positive/<br>Replicates |
| 300 | 0 | 23.9 | 0/3 | 23.9 | 0/3 |
| 300 | 50 | 23.3 | 0/3 | 23 | 0/3 |
| 300 | 100 | 20.5 | 0/3 | 20 | 0/3 |
| 300 | 150 | 19.57 | 0/3 | 19.29 | 0/3 |
| 300 | 200 | 17.41 | 2/3 | 17.81 | 0/3 |
| 300 | 250 | 15.9 | 3/3 | 17.59 | 0/3 |
| 300 | 300 | 15.28 | 3/3 | 14.89 | 3/3 |
| 300 | 400 | 12.99 | 3/3 | 14.19 | 3/3 |
| 300 | 500 | 11.6 | 3/3 | 11.62 | 3/3 |
| 300 | 600 | 10.43 | 3/3 | 11.01 | 3/3 |
| 0 | 300 | 0.1479 | 3/3 | 0.1128 | 3/3 |

**Table 4.** False positives with saline as sample. Results from mixing extraction buffer with varying amounts of 0.9% saline and testing for false positives and conductivity. False positives occurred only when the saline was run without extraction buffer and conductivity was <17.5 mS/cm.

| Extraction Buffer<br>(μL) | Saline<br>(μL) | Lot 2010185/20201307 | | Lot 2010187/20201308 | |
| --- | --- | --- | --- | --- | --- |
|  |  | Conductivity<br>(mS/cm) | False Positive/<br>Replicates | Conductivity<br>(mS/cm) | False Positive/<br>Replicates |
| 300 | 0 | 26.2 | 0/3 | 28.1 | 0/3 |
| 300 | 50 | 23.8 | 0/3 | 24.6 | 0/3 |
| 300 | 100 | 21.9 | 0/3 | 24.2 | 0/3 |
| 300 | 150 | 20.2 | 0/3 | 22.7 | 0/3 |
| 300 | 200 | 20 | 0/3 | 22.3 | 0/3 |
| 300 | 250 | 20 | 0/3 | 21.6 | 0/3 |
| 300 | 300 | 21.6 | 0/3 | 21.9 | 0/3 |
| 300 | 400 | 20 | 0/3 | 22.7 | 0/3 |
| 300 | 500 | 20 | 0/3 | 22.4 | 0/3 |
| 300 | 600 | 19.71 | 0/3 | 21.3 | 0/3 |
| 0 | 300 | 16.18 | 3/3 | 17.25 | 3/3 |

To further support the role of sample conductivity in false-positive results, four VTM solutions were tested for conductivity. Three of the four tested had conductivity of ∼13–15 mS/cm, shown in Table 5, while one of the four had conductivity of ∼ 3 mS/cm. This variability suggests that VTMs can vary in conductivity and may give different results depending on the sample dilution used. Of interest is that the Hardy Diagnostics material with low conductivity also gave one of the false-positive results shown in Table 1.

**Table 5.** UTM/VTM conductivity measurements.

| UTM/VTM | Conductivity (mS/cm) |
| --- | --- |
| Hardy Diagnostics | 2.97 |
| BD Universal Viral Transport Medium | 13.36 |
| COPAN UTM Type 350C | 13.33 |
| XEBIOS Diagnostic GLY 70-2025 | 15.51 |

Viral transport media often contains additives that support viral preservation but could potentially interfere with immunoassays. To investigate this possibility, five of the common VTM components (e.g., bovine serum albumin, fetal bovine serum, gelatin, yeast extract, and albumin) were tested. Solutions of these additives were prepared in assay extraction buffer and tested at levels at or above those observed in VTM. Table 6 shows that all additives generated negative results.

**Table 6.** UTM/ VTM components. All solutions were prepared in extraction buffer and run as normal samples. Negative results were observed for all materials.

| Component | Concentration | Result |
| --- | --- | --- |
| Bovine serum albumin | 100 mg/mL | Negative |
| Fetal bovine serum | 50% (v/v) | Negative |
| Gelatin | 1% (v/v) | Negative |
| Yeast extract | 10% (v/v) | Negative |
| Lactalbumin | 100 mg/mL | Negative |

Laboratory recipes for viral transport media (VTM) are available in the literature and are known to contain 2% fetal bovine serum (FBS) and Hanks balanced salt solution (HBSS) with antimicrobial compounds.^7^ One formulation of a laboratory VTM without antimicrobial compounds was mixed with extraction buffer at several dilutions and tested. Results are shown below in Table 7. All laboratory VTM solutions mixed with extraction buffer up to 1:1 by volume showed negative results. The laboratory VTM run without extraction buffer produced a false-positive result. Conductivity of extraction buffer and VTM are also shown.

**Table 7.**
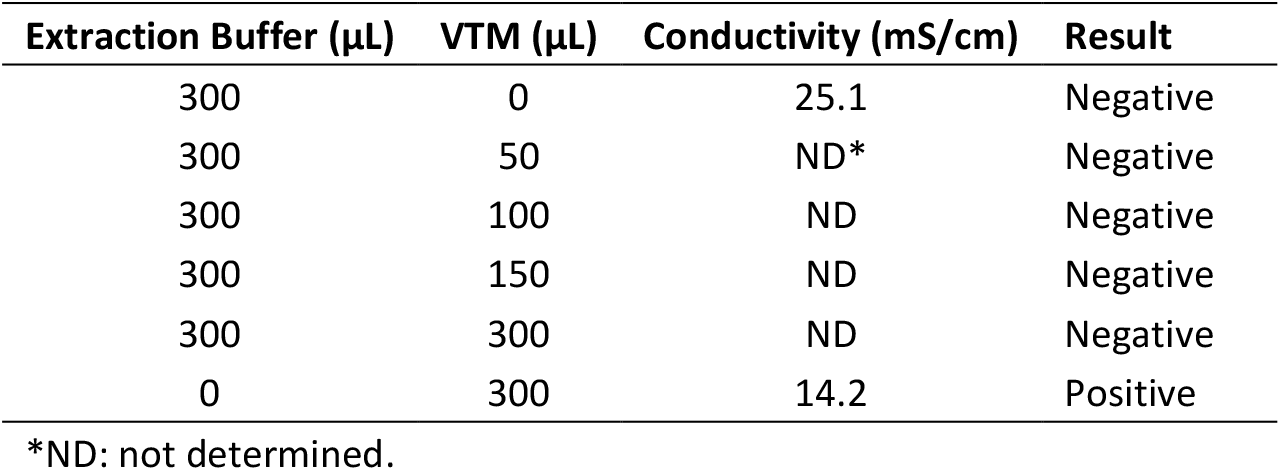
Laboratory VTM. VTM with 2% FBS was mixed with extraction buffer at varying volumes and tested for false-positive results. Conductivity is shown when relevant.

## Discussion

The use of remnant liquid swab samples stored in UTM/VTMs can negatively impact the performance of COVID-19 rapid antigen tests. These sample types are problematic because some manufacturers (e.g., Becton Dickinson, Healgen, and Abbott) do not support the use of UTM/VTMs, others (e.g., AccessBio, SD Biosensor, and Quidel) recommend specific UTM/VTMs, and some specify that certain UTM/VTM products should not be used (e.g., Quidel).^2-5^ UTM/VTM liquid samples are being used in retrospective evaluations of these rapid antigen tests, including the Healgen assay. Some manufacturers use relatively high volumes (e.g., 350–400 µL) of VTM sample to compensate for original sample dilution, thereby improving analytical sensitivity, but this practice could negatively affect the Healgen assay.^3,5^

In fact, during an evaluation of the Healgen rapid antigen test by Charité Hospital, it was noted that the assay exhibited cross-reactivity with seven respiratory non-COVID-19 samples (e.g., adenovirus, entero/rhinovirus, influenza A H1, influenza A H3, and parainfluenza 1, 2, and 3). This was inconsistent with the internal cross-reactivity data generated at Healgen. The authors concluded that one possible cause for the false positives was nonspecific binding. In some cases, duplicate tests of the positive results were performed, which resulted in the expected negative result.^8^

To better understand results generated with UTM/VTM, we screened 12 different materials and found at least 2 that gave a weak false-positive result at 1:1 dilution. Interestingly, none of the materials generated false positives when tested with a 1:6 dilution (i.e., 50 µL of sample:300 µL of extraction buffer). The false-positive results generated with a 1:1 dilution revealed that either dilution of the extraction buffer or material present in the UTM/VTM was the source. Both possibilities were examined thoroughly.

We performed a second study to determine the impact of UTM/VTM liquid samples on the pH, surfactant concentration, and ionic strength. The extraction buffer plays several key roles in assay performance, and when it is mixed with stored liquid samples, the composition can change. The surfactant compounds are key to lysing virus particles (and subsequently inactivating them) and releasing the nucleocapsid protein, making it accessible to the antibodies used in the assay. These compounds also help to reduce nonspecific interactions between the gold sol conjugates and the test and control line antibodies. The pH of the buffer is critical for maintaining a consistent overall charge on all the relevant protein species (e.g., antigen, conjugate, and test and control line antibodies). The ionic strength or salt concentration of the buffer also creates an environment in which nonspecific electrostatic interactions are reduced. Any disruption of these factors may introduce unwanted results. The key finding here is that it appears that ionic strength is the most important factor affecting assay performance with respect to false-positive results using stored samples.

This study was expanded to look at dilution studies with saline and water on two different reagent lots. The water dilution showed the worst-case scenario, as the conductivity of water is very low (C = 0.15 mS/cm). Even at a dilution of 1:1 (water:EB), false-positive results begin to occur. The observed lot differences may be due to slight variation in extraction buffer composition or preparation of the samples tested. Viral samples are not recommended to be stored in water solution for any type of testing; this condition represents an extreme dilution of UTM/VTM samples with water to increase the sample volume for multiple rapid antigen test evaluations and could be problematic. The results of the saline dilution study were more favorable due to the higher conductivity of the 0.9% saline alone (C = 16–17 mS/cm). No false positives were observed when saline was mixed with extraction buffer, even up to 2:1 (saline:EB). Saline is a common way to store viral swabs and provides enough ionic strength to reduce false-positive results. The conductivity measurements from this study have also shown that an approximate cutoff point for false-positive generation with this test is 17 mS/cm.

The findings from the UTM/VTM screening and buffer components study suggested that there may be differences in the conductivity between UTM/VTM from different sources. We measured four materials that have conductivity ranging from about 3–15 mS/cm. This indicates some variability in VTM based on the formula for those materials. The objective of storing swabs in VTM is to preserve and protect the organism for future use, either for cell culture or, most likely in the case of SARS-CoV-2 samples, molecular testing. Different UTM/VTMs have unique purposes, and their formulations reflect those needs. In the case of the Healgen rapid antigen testing, the UTM/VTM with the lowest conductivity caused a weak false-positive result at the 1:1 dilution as expected based on conductivity measurements.

Another possible explanation for poor rapid test performance with UTM/VTM use is that one or more of the components in the UTM/VTMs interfere with the assay. Several different components commonly found in UTM/VTMs were tested at high concentrations (e.g., FCS, BSA, gelatin, lactalbumin, and yeast extract). These components were chosen because they are proteins or contain a mixture of proteins with the potential to interfere in an immunoassay. None of these materials was found to generate false results, further supporting the role of ionic strength in false-positive results from samples stored in UTM/VTM.

We conclude that the most likely reason for discrepant results of the rapid antigen test is the mixture of the UTM/VTM liquid sample with extraction buffer, which may drive nonspecific electrostatic interactions at the assay test line.

## Data Availability

All data available in the manuscript

## Disclaimers

The CLINITEST Rapid COVID-19 Antigen Test is not available for sale in the U.S. Its future availability cannot be guaranteed. Product availability varies by country.

The use of this product with UTM/VTM samples is not an approved sample type as per the IFU for this product as of the date of this publication.

The information in this paper is based on research results that are not commercially available.

CLINITEST and all associated marks are trademarks of Siemens Healthcare Diagnostics Inc., or its affiliates. All other trademarks and brands are the property of their respective owners.

## References

1. Baro B, Rodo P, et al. Performance characteristics of five antigen-detecting rapid diagnostic test (Ag-RDT) for SARS-CoV-2 asymptomatic infection: a head-to-head benchmark comparison. medRxiv 2021.02.11.21251553. doi: https://doi.org/10.1101/2021.02.11.21251553

2. Summary and explanation of the test. BinaxNow COVID-19 Ag Card IFU. Abbott Laboratories.

3. Nasopharyngeal swab in viral transport media (VTM) test procedure. CareStart COVID-19 Antigen IFU. AccessBio.

4. Limitations. Sofia SARS Antigen FIA IFU. Quidel.

5. Specimen collection and preparation. Standard Q COVID-19 Ag Test IFU. SD Biosensor.

6. Kontogianni K, et al. Limit of detection in different matrices of nineteen commercially available rapid antigen tests for the detection of SARS-CoV-2. Available from: https://www.researchsquare.com/article/rs-350333/v1; https://doi.org/10.21203/rs.3.rs-350333/v1

7. Preparation of viral transport medium. SOP# DSR-052-05. Centers for Disease Control and Prevention.

8. Corman VM, Haage VC, et al. Comparison of seven commercial SARS-CoV-2 rapid point-of-care antigen tests. medRxiv 2020.11.12.20230292; doi: https://doi.org/10.1101/2020.11.12.20230292

